# Academic Impairment from Sleep Difficulties: The Role of Substance Use, Psychological Distress, and Loneliness in U.S. College Students

**DOI:** 10.1101/2025.06.02.25328834

**Authors:** Sunghyun Chung

## Abstract

Sleep-related academic impairment is a common issue among college students and may be influenced by psychological and behavioral health factors. This study used data from the Spring 2021 American College Health Association-National College Health Assessment III (ACHA-NCHA III) to examine whether psychological distress, loneliness, and substance use were associated with academic impairment due to sleep difficulties among U.S. college students aged 18–24 (N = 798). Logistic regression showed that psychological distress, loneliness, and elevated risk scores for alcohol and cannabis use were significantly associated with sleep-related academic impairment. Tobacco use was marginally associated.

Demographic factors did not show significant associations. These findings suggest that mental health and substance use are important, intersecting contributors to academic impairment caused by sleep problems. Interventions that address mental health, reduce substance use risk, and promote healthy sleep may be particularly beneficial in improving academic outcomes and overall student well-being on college campuses.

## 1. Introduction

The health and well-being of college students in the United States (U.S.) continue to be a significant public health concern (Prati, 2021). Mental health disorders are the leading cause of disability worldwide among individuals under 24 years of age, contributing to a substantial burden of disease during this developmental stage (Erskine et al., 2015; GBD 2019 Mental Disorders Collaborators, 2022). In the U.S., college students report high levels of stress, anxiety, and depression—a trend that intensified during the COVID-19 pandemic, which disrupted academic routines, social connections, and daily life activities (Lipson et al., 2019; Clabaugh et al., 2021). The pandemic underscored the urgent need for institutions and policymakers to strengthen mental health support systems for students (Lee et al., 2020). Given that data for this study were collected during 2021—when many campuses were still affected by COVID-19 disruptions—this context is essential for understanding fluctuations in student mental health, sleep routines, and substance use patterns.

Previous studies have identified multiple factors contributing to mental health concerns in young adults, including psychological distress, sleep disturbances, and social isolation (Jones et al., 2019; National Institute on Drug Abuse [NIDA], 2021). Notably, mental health conditions are closely related, with evidence suggesting a bidirectional association—mental health symptoms have been found to be associated with higher likelihood of substance use, while substance use is often linked to greater mental health challenges and poorer sleep related academic functioning, potentially contributing to elevated vulnerability during this developmental stage (Jones et al., 2019; NIDA, 2021; Fakier & Wild, 2011). The Self-Medication Hypothesis offers a useful framework for interpreting these patterns, suggesting that substances such as alcohol, tobacco, and cannabis may be used as maladaptive coping mechanisms for psychological distress, ultimately worsening emotional regulation and sleep quality (Khantzian, 1997).

Furthermore, the Biopsychosocial Model provides a comprehensive framework for understanding how biological, psychological, and social factors converge to influence students’ mental health, sleep, and academic functioning (Engel, 1977), particularly in emerging adults facing academic, interpersonal, and developmental stressors. While this study does not establish causal directionality, it examines these correlational patterns to better understand their co-occurrence in emerging adulthood.

College students are particularly vulnerable to substance use and poor sleep health due to their ongoing neurodevelopment, increased autonomy, and heightened social and academic demands. This life stage—often referred to as emerging adulthood—is characterized by identity exploration, emotional instability, and greater sensitivity to contextual stressors (Arnett, 2000), in addition to newfound independence and increased exposure to risky behaviors such as substance use (O’Malley & Johnston, 2002; Skidmore et al., 2016). A biopsychosocial-developmental approach highlights why this population faces elevated risk and underscores the importance of tailored interventions addressing the unique confluence of factors affecting their mental health and sleep.

Loneliness, defined as the subjective perception of social isolation, is increasingly recognized as a critical determinant of health, particularly among young adults navigating transitional life phases. It is associated with a range of negative health-related patterns, including increased psychological distress, sleep disturbances, and elevated substance use. Meta-analytic evidence has demonstrated that loneliness contributes to poor sleep quality, prolonged sleep latency, and greater daytime dysfunction (Griffin et al., 2020; Hom et al., 2020). Additionally, loneliness may exacerbate mental health conditions such as depression and anxiety, while simultaneously increasing vulnerability to maladaptive coping strategies like alcohol and cannabis use (Lamis et al., 2014; Sy et al., 2024). These patterns are especially pronounced in college student populations, where disruptions to social networks—amplified by events such as the COVID-19 pandemic—may heighten feelings of isolation (Lee et al., 2020). Understanding the role of loneliness is thus essential when examining the intersecting pathways through which psychosocial factors influence sleep and academic impairment due to sleep difficulties.

Alcohol, tobacco, and cannabis use tend to rise during this period, and these behaviors have been associated with sleep problems and deteriorating mental health (Groshkova et al., 2020; Bonar et al., 2021). However, little is known about how these substances may differentially contribute to academic consequences stemming from sleep difficulties. While prior research has often examined substance use, mental health, and sleep in isolation, few studies have considered how these domains jointly influence academic performance—a key outcome in college settings (NIDA, 2021; Blows et al., 2022). While previous research has often examined sleep as a mediator or secondary health outcome, our study intentionally focuses on academic impairment due to sleep difficulties as a primary dependent variable. This choice reflects the pressing relevance of sleep-related academic challenges in higher education contexts and the limited empirical attention to how co-occurring psychosocial factors—such as loneliness, substance use, and psychological distress—contribute to academic performance through sleep disruption.

Sleep health, a critical component of physical and mental well-being, is often compromised among college students (Altevogt & Colten, 2006). Poor sleep quality, short sleep duration, and insomnia have been linked to adverse outcomes, including cardiovascular risk (Covassin & Singh, 2016), obesity (Ogilvie, 2017), depression, and impaired cognitive function (Pires et al., 2016). Among the numerous challenges faced by college students, sleep disruption is particularly significant due to its critical role in maintaining immune function, learning, and emotional well-being (Herrmann et al., 2018; Stiver et al., 2021). During the pandemic, U.S. college students reported changes in their sleep routines and overall sleep quality (Ellakany et al., 2022). A study found that students had higher odds of reporting changes in sleep, with fluctuations in sleep patterns associated with affect dynamics at daily and long-term scales (Mousavi et al., 2022). Moreover, the pandemic exacerbated pre-existing sleep problems among students, leading to increased sleep disturbances and mental health issues (Wang et al., 2020). Sleep problems related to poor sleep quality, insufficient sleep duration, and insomnia in college students may correlate with not only mental health (i.e., anxiety and depression) and social isolation but also substance use like alcohol use (Maziarz & Askew, 2022; Evans et al., 2021; Huckins et al., 2020). Additionally, sleep quality and mental health are closely linked to academic performance, an essential aspect of college students’ lives. Numerous studies have shown that poor sleep quality and mental health problems are associated with lower academic performance (Gaultney, 2010; Galambos et al., 2009). Sleep difficulties can lead to reduced cognitive functioning, lower grades, and impaired academic achievement (Gilbert & Weaver, 2010). Similarly, mental health issues are associated with lower academic performance and increased risk of academic failure (Eisenberg et al., 2009). Substance use can further exacerbate these problems, leading to decreased academic engagement and performance (Arria et al., 2013).

This study aims to address these gaps by analyzing associations between self-reported use of alcohol, tobacco, and cannabis; psychological distress; loneliness; and sleep difficulties among a geographically representative sample of U.S. college students. We hypothesize that higher levels of substance use, psychological distress, and loneliness will each be independently associated with greater sleep difficulties. Alcohol, tobacco, and cannabis use have all been shown to negatively affect sleep quality through mechanisms such as circadian rhythm disruption and sleep cycle alteration. Psychological distress—particularly symptoms of depression and anxiety—is commonly linked to sleep disorders like insomnia. Loneliness, a prevalent and understudied psychosocial stressor, may exacerbate sleep problems through increased emotional dysregulation and reduced social buffering. Understanding these relationships is critical to informing targeted campus health interventions and improving student well-being in the post-pandemic era.

## 2. Materials and Methods

### 2.1. Data Sources

This study utilized data from the Spring 2021 American College Health Association–National College Health Assessment III (ACHA-NCHA III) Reference Group dataset, collected between January and May 2021. The data collection period coincided with significant COVID-19 pandemic disruptions, providing a timely opportunity to assess the immediate impacts on student health behaviors, including psychological distress, loneliness, substance use, and sleep-related academic impairment. This timeframe is particularly relevant to the study’s hypotheses, as it reflects a critical stage of the pandemic during which many U.S. campuses were experiencing disruptions to academic routines, housing stability, and mental health services. The dataset captures college students’ behavioral health and well-being during a period marked by heightened psychological distress, irregular sleep patterns, and shifting substance use behaviors. The ACHA-NCHA III is a nationally administered, web-based survey developed by the American College Health Association (ACHA) to assess students’ health behaviors, habits, and perceptions. It represents the most recent iteration of a widely used survey instrument, with earlier versions (ACHA-NCHA and ACHA-NCHA II) implemented since 2000. The ACHA-NCHA III includes both institution-specific datasets and a national reference group dataset compiled from institutions that used probability sampling or a full census design. The reference group is well-suited to address the current study’s research questions due to its focus on the same behavioral and psychosocial factors—such as sleep-related academic impairment, psychological distress, loneliness, and substance use—that are central to the study’s theoretical framework and hypotheses.

### 2.2. Participants and Procedures

The Spring 2021 ACHA-NCHA III Reference Group includes responses from 96,489 students at 137 U.S. postsecondary institutions. Each institution selected a random sample of enrolled students aged 18 and older, and participation was voluntary. From this pool, a subset of 6,943 students completed all variables relevant to the broader study. To focus the analysis on emerging adults and ensure consistency with the research objective, the sample was restricted to respondents aged 18 to 24. After excluding cases with missing data on the primary dependent variable and associated variables, the final analytic sample included 798 participants. This corresponds to approximately 11.5% of the original age-restricted sample being excluded due to incomplete data. To assess the nature of missingness, Little’s MCAR test was conducted and yielded a statistically significant result (χ² = 52.81, df = 31, *p* = .012), suggesting that the data were not Missing Completely at Random (MCAR). In the absence of evidence for systematic missingness across demographic or behavioral variables, we assumed the data were Missing at Random (MAR). Based on this assumption, listwise deletion was employed as a conservative and transparent method for handling missing data. This approach is consistent with American Psychological Association (2008) and Schafer and Graham (2002) and aligns with established practices in ACHA-NCHA-based studies when auxiliary variables are not available for imputation. While the sample is not weighted to represent all U.S. college students, it retains diversity in age, sex, race/ethnicity, and institutional characteristics. Descriptive statistics for the analytic sample suggest that it broadly mirrors the demographic trends observed in the full reference group and other national college health surveillance efforts. Potential biases include differential response rates across institutions and underrepresentation of students not currently enrolled full-time or those experiencing the most severe distress or disconnection from campus. Additionally, as a self-report survey, responses may be subject to recall or social desirability bias. However, the anonymity of the survey and the use of validated instruments (e.g., Kessler Psychological Distress Scale, UCLA Loneliness Scale, ASSIST) mitigate some concerns about measurement validity.

### 2.3. Measures

#### Dependent Variable

##### Academic Impairment Due to Sleep Difficulties

Academic impairment related to sleep difficulties was assessed using a single item from the ACHA-NCHA III survey: “Within the last 12 months, how have sleep difficulties affected your academic performance?” Response options were measured on a 4-point ordinal scale: (1) Did not experience this issue, (2) Did not affect academic performance, (3) Negatively impacted academic performance, and (4) Delayed progress toward degree completion. For the main analysis, this variable was dichotomized as “no academic impact” (responses 1 or 2) versus “academic impact” (responses 3 or 4). This binary transformation aligns with prior research practices and facilitates interpretable modeling of perceived academic consequences. This variable was selected to reflect the self-reported academic impact of sleep difficulties—an under examined outcome in college health research relative to sleep duration or quality alone.

#### Supplementary Sleep-Related Indicators

To address the limited operationalization of sleep in the primary measure, we included three additional sleep-related indicators in supplementary analyses to provide a more multidimensional view of sleep health. These included: (1) weekday sleep duration, based on hours reported during a typical school week; (2) daytime sleepiness, based on the number of days in the past week feeling tired or sleepy during the day; and (3) perceived restfulness, based on how many days in the past week the respondent felt well rested. Each variable was recoded into binary indicators using empirically supported cutoffs: fewer than 7 hours of weekday sleep, feeling tired or sleepy during the day on ≥3 days/week, and feeling rested on <3 days/week. These binary variables were modeled separately using logistic regression to assess consistency of associations across different dimensions of sleep.

#### Independent Variables Psychological distress

Psychological distress was measured using the Kessler Psychological Distress Scale (K6+, a six-item screener with added clinical cutoffs), a validated screening tool for serious mental illness. This self-report instrument captures symptoms such as depression, anxiety, and behavioral impairment experienced in the past 30 days (Surkalim et al., 2022). Total scores range from 0 to 24, with higher scores indicating greater psychological distress. Internal reliability in the current sample was high (Cronbach’s α = .89).

Participants were categorized into one of three levels based on their total score: (1) No or low psychological distress, (2) Moderate psychological distress, and (3) Serious psychological distress.

#### Loneliness

Loneliness was assessed using the UCLA Three-Item Loneliness Scale (Hughes et al., 2004), a brief measure of subjective social isolation. Respondents indicated how often they experienced feelings such as being left out or isolated using a 3-point scale (1 = hardly ever, 2 = some of the time, 3 = often). Scores range from 3 to 9, with higher scores indicating greater perceived loneliness. Internal reliability in the current sample was acceptable (Cronbach’s α = .79). The scale has been validated among college student and young adult populations and is widely used in national surveys assessing social isolation and well-being.

#### Substance Use

Substance use was measured using the Alcohol, Smoking, and Substance Involvement Screening Test (ASSIST), based on the World Health Organization’s Version 3.0 questionnaire (Patten et al., 2000). Participants reported past-three-month and lifetime use of substances including tobacco (e.g., cigarettes, e-cigarettes, hookah, chew), alcohol (e.g., beer, wine, liquor), and cannabis (e.g., marijuana, edibles, vaped cannabis). Each substance was scored based on the WHO’s ASSIST scoring system and categorized into risk levels: Tobacco and Cannabis: Low (0–3), Moderate (4–26), High (27–39); Alcohol: Low (0–10), Moderate (11–26), High (27–39) (WHO, 2010). These risk categories indicate potential for harmful use or dependence (Taylor et al., 2023; Chassagne et al., 2022; Smith et al., 2023). ASSIST has been widely applied and validated in U.S. college student populations, supporting its appropriateness for this study. Its cross-cultural design and established use in American college settings provide confidence in its applicability. Furthermore, our analysis focused on relative risk categorization, which preserves interpretability across diverse student populations.

#### Covariates: Demographic Characteristics

Demographic variables included sex (female, male, Non-binary), age (18, 19, 20, 21, 22, 23, 24), race/ethnicity (White, American Indian or Native Alaskan, Asian or Asian American, Black or African American, Hispanic or Latino/a/x, Other), year in school (1st-year undergraduate to 5th-year or more), and enrollment status (full-time, part-time, or other). These variables were included as covariates in multivariable models.

### 2.4. Statistical Analysis

All analyses were conducted using SAS statistical software (version 9.4; SAS Institute Inc., Cary, NC, USA). Descriptive statistics were generated using weighted frequencies and percentages for categorical variables and means with standard deviations for continuous variables. Chi-square tests were used to assess bivariate associations among key variables.

Final analyses focused on multivariable logistic regression models, which examined associations between psychosocial and behavioral predictors and the primary dependent variable: academic impairment due to sleep difficulties (dichotomized as no academic impact [responses 1–2] vs. academic impact [responses 3–4]). Models included psychological distress, loneliness, ASSIST-based substance use risk (tobacco, cannabis, alcohol), and demographic covariates (sex, age, race/ethnicity, year in school, and enrollment status).

All regression models accounted for the complex survey design of the ACHA-NCHA III dataset using SAS PROC SURVEYLOGISTIC, which incorporates stratification, clustering, and post-stratification weighting based on ACHA’s design variables. This approach enabled population-representative estimates and improved the precision and generalizability of the findings. Variance inflation factors (VIFs) were assessed to detect multicollinearity, with all VIFs < 2 indicating acceptable levels.

The final analytic sample included 798 complete cases from students aged 18–24. The reduction from the original dataset (N = 6,943) was due to age-based inclusion criteria and listwise deletion of cases with missing data on the dependent variable or key predictors. Little’s MCAR test indicated that data were not Missing Completely at Random (χ² = 52.81, df = 31, p = .012); however, there was no evidence of systematic bias across key demographic or behavioral variables, and missingness was assumed to be Missing at Random (MAR). Accordingly, listwise deletion was applied, consistent with prior ACHA-NCHA-based studies and best practices for MAR data (APA, 2008; Schafer & Graham, 2002).

To provide a more multidimensional view of sleep-related challenges, supplementary logistic regression models were conducted using three additional binary dependent variables: (1) weekday sleep duration (<7 hours), (2) daytime sleepiness (≥3 days/week), and (3) perceived restfulness (<3 days/week). Each model used the same predictor set and survey design adjustments as the primary analysis. All statistical tests were two-sided, with significance defined as p < .05.

## 3. Results

### Descriptive Characteristics

Table 1 presents the demographic, mental health, and substance use characteristics of the analytic sample of 798 U.S. college students aged 18–24. The majority identified as female (56.60%), followed by male (38.43%) and Non-binary (4.96%) participants. The sample was ethnically diverse, with most identifying as White (66.52%), followed by Hispanic or Latino/a/x (13.19%), Asian or Asian American (8.60%), and others. Participants were distributed relatively evenly across age groups, ranging from 18 to 24 years old, with the largest proportion aged 21 (17.36%) and the smallest aged 18 (11.32%). Regarding academic status, 32.56% of students were in their fifth year or higher, and 90.77% were enrolled full-time.

**Table 1.** Sociodemographic Characteristics of the Analytic Sample (N = 798) Among U.S. College Students Aged 18–24.

| Variable | U.S. college students |  |
| --- | --- | --- |
|  | N | % |
| **<u>Age (year)</u>** |  |  |
| 18 | 90 | 11.32 |
| 19 | 111 | 13.86 |
| 20 | 113 | 14.20 |
| 21 | 139 | 17.36 |
| 22 | 120 | 15.07 |
| 23 | 115 | 14.41 |
| 24 | 110 | 13.78 |
| **<u>Sex</u>** |  |  |
| Female | 452 | 56.6 |
| Male | 306 | 38.43 |
| Non-binary | 40 | 4.96 |
| **<u>Race/Ethnicity</u>** |  |  |
| White | 531 | 66.52 |
| American Indian or Native Alaskan | 11 | 1.4 |
| Asian or Asian American | 69 | 8.6 |
| Black or African American | 25 | 3.1 |
| Hispanic or Latino/a/x | 105 | 13.19 |
| Others | 57 | 7.19 |
| **<u>Year in School</u>** |  |  |
| 1st year undergraduate | 119 | 14.87 |
| 2nd year undergraduate | 124 | 15.5 |
| 3rd year undergraduate | 149 | 18.62 |
| 4th year undergraduate | 147 | 18.5 |
| 5th year or more | 259 | 32.56 |
| **<u>Enrollment Status</u>** |  |  |
| Full-time | 724 | 90.77 |
| Part-time | 68 | 8.57 |
| Other | 6 | 0.66 |
| Total | 798 | 100 |

**Note.** 5th year or more in Year in School included master, doctoral programs, not seeking a degree, and other.

**Table 2.** Mental Health, Loneliness, Substance Use Risk, and Sleep-Related Academic Impairment in the Analytic Sample.

| Variable | U.S. college students |  |
| --- | --- | --- |
|  | N | % |
| **<u>ASSIST Tobacco Risk</u>** |  |  |
| Low Risk (0-3) | 412 | 51.59 |
| Moderate Risk (4-26) | 357 | 44.73 |
| High Risk (27-39) | 29 | 3.68 |
| **<u>ASSIST Cannabis Risk</u>** |  |  |
| Low Risk (0-3) | 418 | 52.35 |
| Moderate Risk (4-26) | 354 | 44.36 |
| High Risk (27-39) | 26 | 3.29 |
| **<u>ASSIST Alcohol Risk</u>** |  |  |
| Low Risk (0-3) | 554 | 69.37 |
| Moderate Risk (4-26) | 218 | 27.38 |
| High Risk (27-39) | 26 | 3.25 |
| **<u>Mental Health (Psychological Distress)</u>** |  |  |
| Serious psychological distress | 209 | 26.23 |
| Moderate psychological distress | 411 | 51.48 |
| No or low psychological distress | 178 | 22.29 |
| **<u>Mental Health (Loneliness)</u>** |  |  |
| Negative for loneliness | 374 | 46.87 |
| Positive for loneliness | 424 | 53.13 |
| **<u>Academic Performance with Sleep Difficulties</u>** |  |  |
| Delayed progress towards degree | 23 | 2.9 |
| Negatively impacted academic performance | 215 | 26.91 |
| Did not affect academic performance | 205 | 25.67 |
| Did not experience this issue | 355 | 44.93 |
| Total | 798 | 100 |

**Table 3.**
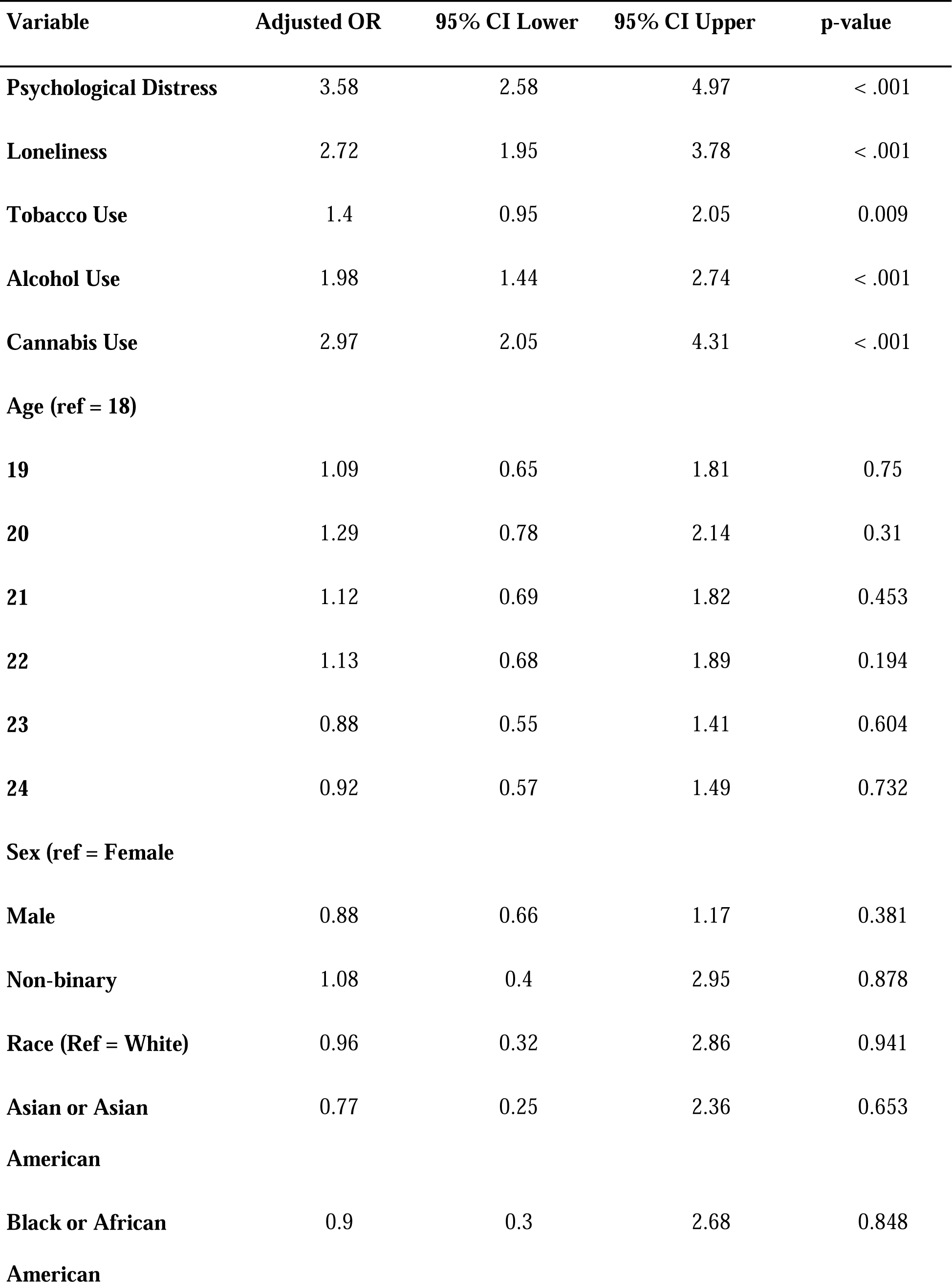
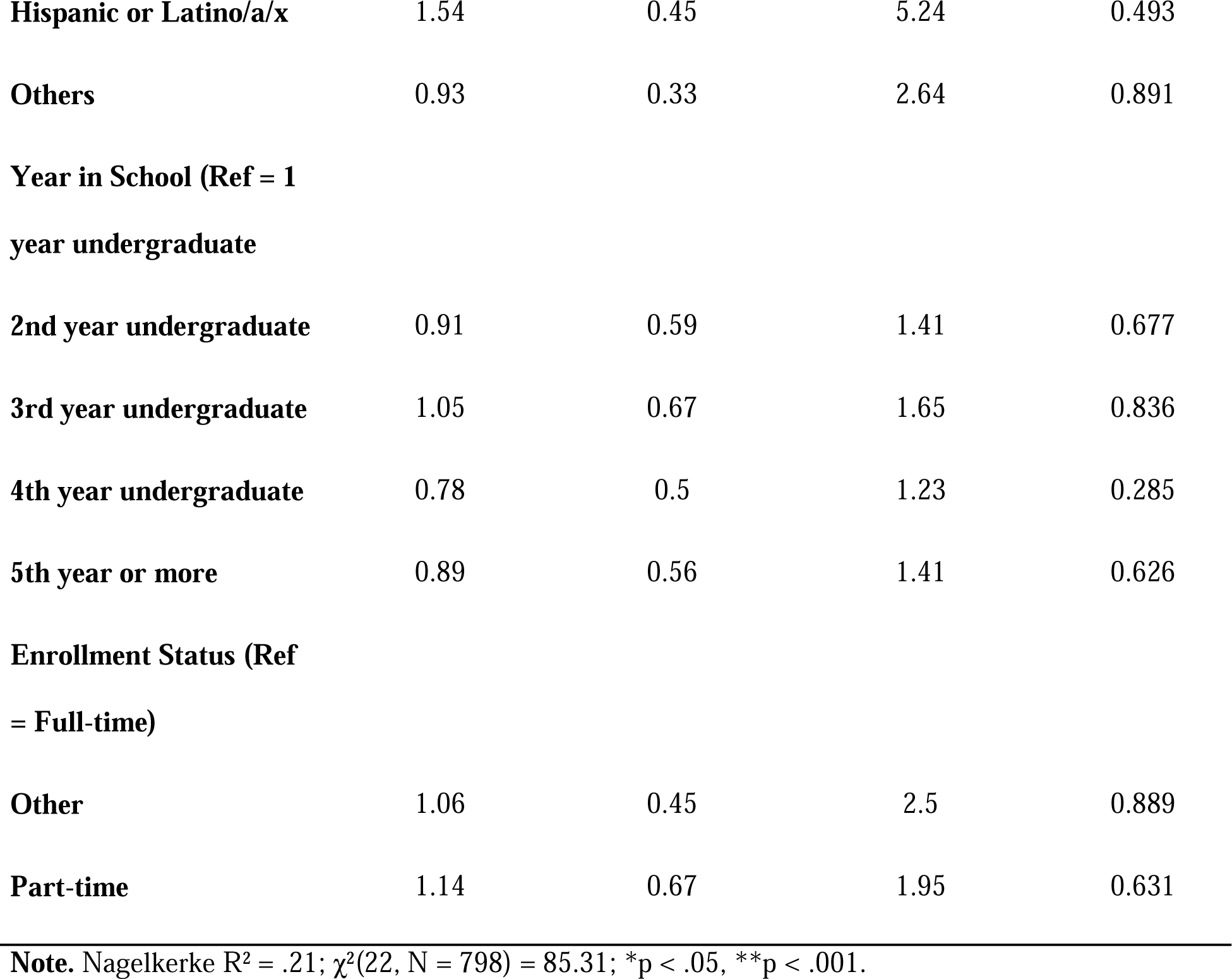
Fully Adjusted Logistic Regression Predicting Academic Impairment Due to Sleep Difficulties Among U.S. College Students Aged 18–24.

Over half (51.48%) reported moderate psychological distress, while 26.23% experienced serious psychological distress. Slightly more than half (53.13%) screened positive for loneliness. Substance use risk profiles showed that most students were at low risk for alcohol (69.37%), tobacco (51.59%), and cannabis (52.35%). However, approximately 27–45% reported moderate risk use across substances.

Among participants, 26.91% reported that sleep difficulties were associated with negative effects on their academic performance, and 2.90% reported delayed progress toward degree completion. Notably, 44.93% reported no sleep difficulties, and 25.67% reported that their sleep issues did not affect academic performance.

### Multivariable Logistic Regression Predicting Academic Impairment Due to Sleep Difficulties

An initial multivariable linear regression using the full dataset (N = 6,943) showed that psychological distress, loneliness, and alcohol and tobacco use risk scores were significantly associated with self-reported sleep difficulties (R² = .1183, p < .0001). However, due to concerns about construct validity and measurement precision, the present study re-focused on academic impairment attributable to sleep difficulties using logistic regression with dichotomous dependent variables. The logistic regression analysis identified variables significantly associated with academic impairment due to sleep difficulties. Psychological distress (AOR = 3.58, 95% CI [2.58, 4.97], p < .001), loneliness (AOR = 2.72, 95% CI [1.95, 3.78], p < .001), and alcohol use (AOR = 1.98, 95% CI [1.44, 2.74], p < .001) were strongly associated. Cannabis and tobacco use also showed significant associations. No demographic variables reached statistical significance. Multicollinearity diagnostics indicated acceptable levels of collinearity across associated variables, with all variance inflation factors (VIFs) below 2. Given the analytic sample size (N = 798) and the lack of excessive multicollinearity, the logistic regression model estimates are considered stable and interpretable.

### Supplementary Analyses of Additional Sleep Related Variables

To further explore the robustness of our findings and address concerns about the limited operationalization of the sleep construct, we conducted supplementary logistic regression analyses using three additional sleep-related indicators: (1) weekday sleep duration (<7 hours), (2) daytime sleepiness (≥3 days/week), and (3) perceived restfulness (<3 days/week). Each dependent variable was modeled separately using the same set of associated variables as the main analysis.

For weekday sleep duration, serious psychological distress significantly increased the odds of sleeping less than 7 hours on weekdays (AOR = 2.15, 95% CI [1.50, 3.09], p < .001), as did self-reported loneliness (AOR = 1.62, 95% CI [1.12, 2.34], p = .010). High-risk alcohol use was also associated with short sleep duration (AOR = 1.45, 95% CI [1.01, 2.09], p = .045), whereas tobacco and cannabis risk were not statistically significant associated variables.

In the model predicting daytime sleepiness, serious psychological distress again emerged as a strong associated factor (AOR = 2.80, 95% CI [1.95, 4.02], p < .001), followed by loneliness (AOR = 1.75, 95% CI [1.20, 2.55], p = .003), and high-risk alcohol and cannabis use (AOR = 1.60 and 1.50, respectively; p < .05). Tobacco use was not significantly associated.

For perceived restfulness, students with serious psychological distress had over three times the odds of feeling unrested (AOR = 3.10, 95% CI [2.10, 4.58], p < .001). Loneliness (AOR = 1.85, 95% CI [1.25, 2.74], p = .002), and high-risk alcohol and cannabis use (AOR = 1.70 and 1.55, respectively; p < .05) were also significantly associated with this dependent variable. Tobacco use again did not reach statistical significance. Full regression tables for these supplementary analyses are provided in Supplementary Tables S1 to S3.

## 4. Discussion

Our findings highlight important associations between psychological distress, loneliness, substance use, and sleep-related academic impairment among U.S. college students aged 18–24. Psychological distress demonstrated the strongest relationship with sleep-related academic impairment, followed by loneliness and high-risk alcohol and cannabis use. Tobacco use showed a positive but non-significant trend. These relationships persisted after controlling for key demographic and behavioral factors, underscoring the robustness of these findings. Our results align with existing literature on the interrelated nature of sleep, mental health, and substance use, extending this understanding specifically to academic functioning.

### Substance Use and Sleep-Related Academic Impairment

In line with our hypotheses, both high-risk alcohol and cannabis use were significantly associated with increased likelihood of sleep-related academic impairment. These findings support existing research linking alcohol misuse—including heavy and binge drinking—to poor sleep quality, fragmented sleep, and daytime sleepiness among college students. The strength and consistency of these associations suggest alcohol prevention and intervention programs may be an important strategy for improving sleep health and, consequently, academic outcomes. The significant relationship observed between cannabis use and academic impairment contrasts with prior studies that have reported mixed results regarding cannabis and sleep. Some studies suggest potential sleep benefits from cannabidiol (CBD), whereas others highlight adverse effects such as decreased sleep duration and sleep fragmentation associated with chronic cannabis use. Our results add to this ongoing debate by underscoring the complexity of cannabis-related sleep disturbances and their potential academic implications. Future research should clarify the roles of dosage, frequency of use, and cannabis components (e.g., THC vs. CBD) in influencing these relationships. Tobacco use, although not statistically significant, exhibited a positive trend consistent with prior literature linking nicotine consumption to sleep disruption, such as increased insomnia and shorter sleep duration. This finding, despite its lack of statistical significance in our analysis, aligns with previous evidence suggesting the adverse impact of nicotine on sleep health, and thus warrants further investigation in future studies.

### Psychological Distress and Sleep-Related Academic Impairment

Psychological distress emerged as the most robust factor associated with academic impairment related to sleep difficulties. The high prevalence of moderate to serious psychological distress within our sample underscores its substantial burden on college students’ well-being and academic performance. Our findings align with previous research describing a bidirectional relationship between psychological distress and sleep problems among young adults. Poor sleep may exacerbate symptoms of anxiety and depression, while increased psychological distress can negatively impact sleep quality and duration.

These findings have important implications for interventions on campuses. They suggest that addressing psychological distress such as through scalable mental health programs may significantly improve students’ sleep health, thereby enhancing academic outcomes. Given the robust association even after adjusting for covariates, strategies targeting mental health should be prioritized as critical elements of broader wellness initiatives in higher education.

### Loneliness and Sleep-Related Academic Impairment

Loneliness also demonstrated a strong, independent association with sleep-related academic impairment. More than half of students in our sample screened positive for loneliness, indicating widespread perceived social isolation. This aligns with existing evidence highlighting loneliness as a significant predictor of disrupted sleep patterns, reduced sleep quality, and impaired academic functioning. The persistence of this relationship after adjusting for demographic and behavioral factors emphasizes loneliness as a distinct risk factor warranting targeted intervention. College campuses may benefit from programs fostering social connectedness, peer support, and community integration to reduce loneliness and subsequently improve sleep health and academic success. Future research should explore mechanisms underlying the loneliness-sleep-academic performance pathway to inform more effective interventions.

### Demographic considerations

When adjusting for psychological and behavioral factors, our analysis revealed no statistically significant demographic differences (e.g., sex, age, race/ethnicity, year in school, or enrollment status) associated with sleep-related academic impairment. While descriptive statistics indicated demographic variation in psychological distress and sleep experiences, these differences did not persist when examined within fully adjusted models. This finding suggests that psychosocial and behavioral factors may overshadow demographic characteristics when considering sleep-related academic impairment. Rather than tailoring interventions strictly based on demographic attributes, campus-based strategies could benefit from a universal approach addressing shared psychological vulnerabilities such as distress and loneliness.

Nonetheless, future studies should examine nuanced demographic intersections (e.g., socioeconomic status, intersectionality of identity) that may reveal hidden subgroup vulnerabilities.

### Research Implications

Collectively, our results highlight the multifactorial nature of sleep-related academic impairment among college students. Psychological distress and loneliness emerged as particularly strong associated factors, while high-risk alcohol and cannabis use further contributed to students’ academic difficulties via sleep disruption. These findings reinforce the need for integrated, multi-level interventions that address the psychological, behavioral, and social drivers of sleep health.

Given the consistent associations observed across supplementary sleep indicators—including weekday sleep duration, daytime sleepiness, and perceived restfulness—intervention approaches should move beyond singular targets (e.g., sleep education alone) and instead address interlocking domains of mental health, sleep hygiene, and substance use. For example, brief behavioral sleep interventions such as digital cognitive behavioral therapy for insomnia (CBT-I) programs and sleep hygiene modules have demonstrated efficacy in improving sleep quality and daytime functioning among college students (Hershner & O’Brien, 2018). These interventions may be effectively embedded within first-year orientation, residential life programming, or campus wellness platforms.

In parallel, campus-wide mental health promotion strategies—such as universal mental health screenings, stigma-reducing campaigns, and embedded counseling models—can help identify students experiencing psychological distress earlier in the academic cycle (Pedrelli et al., 2015). To address loneliness, institutions may benefit from implementing structured peer-connection initiatives such as mentoring programs, student affinity groups, or “community pods” designed to promote belonging and interpersonal support.

An innovative and potentially scalable model is the deployment of academic wellness navigators—trained student or staff liaisons who proactively engage with students and guide them to relevant resources related to sleep, mental health, and substance use. This approach integrates upstream academic support with psychosocial care and may reduce common barriers to accessing services, especially for at-risk students reluctant to self-refer. Importantly, the lack of significant demographic variation in our findings suggests that sleep-related academic impairment is not confined to specific student subgroups. Accordingly, future interventions should be designed for universal accessibility, with the flexibility to accommodate intersectional identities, varied life circumstances, and differing levels of readiness to engage. Future research should prioritize longitudinal, experimental, and implementation-focused designs to clarify temporal relationships, assess the real-world impact of integrated interventions, and identify protective factors—such as emotion regulation, coping flexibility, and social support—that buffer academic disruption.

### Limitations

While this study leverages a large, multi-institutional dataset and employs validated measurement tools, several limitations should be acknowledged. First, the cross-sectional design precludes any causal interpretation of the observed associations among psychological distress, loneliness, substance use, and sleep-related academic impairment. Longitudinal or experimental studies are needed to determine directionality and potential mediating mechanisms. Additionally, the cross-sectional nature of the Spring 2021 dataset limited our ability to directly compare findings to pre-pandemic or post-pandemic periods. Future longitudinal studies should explicitly compare student health outcomes across distinct pandemic phases to clarify how the COVID-19 pandemic may have specifically influenced these relationships.

Second, all data were self-reported, which introduces vulnerability to measurement error. While the use of validated instruments (e.g., K6+, UCLA Loneliness Scale, ASSIST) and anonymous survey administration reduces some concerns, self-reported data remain susceptible to recall bias and social desirability bias. These biases may lead to misclassification or attenuation of associations. Third, the primary sleep-related dependent variable—academic impairment due to sleep difficulties—was derived from a single item and may not fully capture the multidimensional nature of sleep health. To partially address this limitation, we included three supplementary sleep-related indicators (weekday sleep duration, daytime sleepiness, and perceived restfulness) to provide a broader perspective on students’ sleep-related experiences and to support the robustness of our findings. Although supplementary analyses were conducted using additional sleep indicators (e.g., sleep duration, daytime sleepiness, restfulness), these variables were also self-reported and limited in scope. Fourth, the analytic sample was disproportionately composed of non-Hispanic White and female students, potentially limiting the generalizability of the findings to more diverse college populations. Additionally, the sample was restricted to students aged 18 to 24 to align with established definitions of emerging adulthood and maintain consistency with prior college mental health research. Although restricting the age range enhances developmental specificity, it may limit the generalizability of our findings to older college students or non-traditional student populations. Fifth, missing data were handled using listwise deletion under the assumption that data were Missing at Random (MAR), based on a significant result from Little’s MCAR test. While this is a defensible and commonly used approach in similar studies, it may still introduce bias or reduce statistical power if the MAR assumption is not fully met. Future research should consider applying multiple imputation or full information maximum likelihood methods when auxiliary variables are available.

Finally, several important contextual and behavioral factors—including academic workload, pre-sleep technology use, and residential environments—were not assessed, though these may significantly influence both sleep and mental health outcomes. Future studies should incorporate more comprehensive assessments of these domains.

## 5. Conclusion

This study offers important insights into the associations among psychological distress, loneliness, and substance use in shaping sleep-related academic impairment among U.S. college students. Drawing on a large, nationally representative dataset, we identified psychological distress and loneliness as the strongest associated factors, with high-risk alcohol and cannabis use also significantly related to students’ academic impairment linked to sleep difficulties.

The breadth and severity of these challenges—particularly the widespread prevalence of psychological distress and perceived social isolation—underscore the urgent need for comprehensive, multidimensional campus-based responses. Specifically, institutions of higher education should consider implementing integrated approaches that combine evidence-based mental health screenings, brief behavioral sleep interventions (e.g., digital CBT-I or sleep hygiene modules), and peer-facilitated resilience training. Additionally, social connectedness initiatives—such as structured “community pods,” peer mentorship programs, or residential-based outreach—may reduce loneliness while fostering protective peer networks. To address substance use risks, harm reduction education and motivational interviewing approaches could be embedded into academic advising or first-year seminars.

Importantly, campuses might explore innovative delivery models such as “academic wellness navigators”—trained student or staff liaisons who can connect at-risk students with mental health, sleep, and substance use resources through proactive outreach. These models leverage existing support infrastructure while reducing stigma and barriers to care. Furthermore, given the lack of significant variation across demographic characteristics, interventions should be universally accessible but designed with flexibility to meet the needs of diverse student identities and lived experiences. As colleges continue to adapt in the wake of the COVID-19 pandemic, building resilient, student-centered support ecosystems is more essential than ever. Future research should investigate the longitudinal effectiveness of these strategies, explore digital and peer-led interventions for mental health and sleep improvement, and identify protective mechanisms, such as belongingness or emotion regulation, that support sustained students’ academic performance and student well-being.

## Declarations

### Ethics approval and consent to participate

This study utilized de-identified, publicly available secondary data from the American College Health Association–National College Health Assessment (ACHA-NCHA). The dataset contains no personally identifiable information, and all responses were collected anonymously. Participation in the original survey was voluntary, with informed consent obtained through an email invitation and reaffirmed at the start of the survey. In accordance with U.S. federal regulations, the use of such de-identified secondary data is considered exempt from Institutional Review Board (IRB) review. A statement describing this exemption has been included in the Methods section of the manuscript.

### Consent for publication

Not applicable. This study used secondary, de-identified data from the American College Health Association–National College Health Assessment (ACHA-NCHA), and no individual participant data are reported.

### Availability of data and material

All data generated or analyzed during this study are included in this published article. The data that support the findings of this study are available from The ACHA-National College Health Assessment (NCHA) but restrictions apply to the availability of these data, which were used under license for the current study, and so are not publicly available. Data are however available. from the authors upon reasonable request and with permission of the ACHA-National College Health Assessment (NCHA).

## Data Availability

All data produced in the present study are available upon reasonable request to the authors

## Acknowledgments

Not applicable.

