## Supplemental Tables for "Academic Impairment from Sleep Difficulties: The Role of Substance Use, Psychological Distress, and Loneliness in U.S. College Students"

Supplementary Table
**Table S1**

Fully Adjusted Logistic Regression Predicting Short Weekday Sleep Duration (<7 Hours) Among U.S. College Students Aged 18–24 (N = 798)

| **Variable** | **Adjusted OR** | **95% CI Lower** | **95% CI Upper** | **p-value** |
| --- | --- | --- | --- | --- |
| **Psychological Distress** | 2.15 | 1.50 | 3.09 | < .001** |
| **Loneliness** | 1.62 | 1.12 | 2.34 | 0.01* |
| **Tobacco Use** | 1.25 | 0.88 | 1.78 | 0.2 |
| **Alcohol Use** | 1.45 | 1.01 | 2.09 | .045* |
| **Cannabis Use** | 1.38 | 0.95 | 2.00 | 0.085 |
| **Age (ref = 18)** |  |  |  |  |
| **19** | 1.05 | 0.65 | 1.70 | 0.85 |
| **20** | 0.98 | 0.61 | 1.58 | 0.94 |
| **21** | 0.95 | 0.59 | 1.54 | 0.85 |
| **22** | 0.92 | 0.57 | 1.49 | 0.732 |
| **23** | 0.88 | 0.55 | 1.41 | 0.604 |
| **24** | 0.92 | 0.57 | 1.49 | 0.732 |
| **Sex (ref = Female** |  |  |  |  |
| **Male** | 1.08 | 0.76 | 1.54 | 0.66 |
| **Non-binary** | 1.20 | 0.66 | 2.17 | 0.55 |
| **Race (Ref = White)** |  |  |  |  |
| **Asian or Asian American** | 0.91 | 0.58 | 1.44 | 0.69 |
| **Black or African American** | 0.94 | 0.55 | 1.62 | 0.82 |
| **Hispanic or Latino/a/x** | 1.10 | 0.73 | 1.64 | 0.64 |
| **Others** | 0.93 | 0.33 | 2.64 | 0.891 |
| **Year in School (Ref = 1 year undergraduate** |  |  |  |  |
| **2nd year undergraduate** | 0.93 | 0.61 | 1.42 | 0.74 |
| **3rd year undergraduate** | 1.05 | 0.69 | 1.60 | 0.82 |
| **4th year undergraduate** | 1.01 | 0.67 | 1.52 | 0.98 |
| **5th year or more** | 0.97 | 0.63 | 1.49 | 0.89 |
| **Enrollment Status (Ref = Full-time)** |  |  |  |  |
| **Other** | 1.08 | 0.61 | 1.90 | 0.79 |
| **Part-time** | 0.90 | 0.47 | 1.74 | 0.76 |

**Note.** Nagelkerke R² = .18; χ²(22, N = 798) = 71.02; *p < .05, **p < .001.

Table S2

Fully Adjusted Logistic Regression Predicting Daytime Sleepiness (≥3 Days per Week) Among U.S. College Students Aged 18–24 (N = 798)

| **Variable** | **Adjusted OR** | **95% CI Lower** | **95% CI Upper** | **p-value** |
| --- | --- | --- | --- | --- |
| **Psychological Distress** | 2.80 | 1.95 | 4.02 | <.001** |
| **Loneliness** | 1.75 | 1.20 | 2.55 | .003* |
| **Tobacco Use** | 1.35 | 0.95 | 1.92 | .090 |
| **Alcohol Use** | 1.60 | 1.10 | 2.32 | .015* |
| **Cannabis Use** | 1.50 | 1.05 | 2.15 | .030* |
| **Age (ref = 18)** |  |  |  |  |
| **19** | 0.98 | 0.67 | 1.65 | 0.857 |
| **20** | 0.93 | 0.63 | 1.57 | 0.946 |
| **21** | 0.84 | 0.49 | 1.51 | 0.853 |
| **22** | 0.92 | 0.59 | 1.38 | 0.72 |
| **23** | 0.97 | 0.47 | 1.39 | 0.588 |
| **24** | 0.92 | 0.59 | 1.40 | 0.736 |
| **Sex (ref = Female** |  |  |  |  |
| **Male** | 1.17 | 0.79 | 1.58 | 0.669 |
| **Non-binary** | 1.15 | 0.67 | 2.25 | 0.541 |
| **Race (Ref = White)** |  |  |  |  |
| **Asian or Asian American** | 0.90 | 0.49 | 1.50 | 0.679 |
| **Black or African American** | 0.91 | 0.45 | 1.58 | 0.831 |
| **Hispanic or Latino/a/x** | 1.10 | 0.78 | 1.61 | 0.632 |
| **Others** | 1.10 | 0.51 | 1.73 | 0.943 |
| **Year in School (Ref = 1 year undergraduate** |  |  |  |  |
| **2nd year undergraduate** | 1.02 | 0.60 | 1.41 | 0.75 |
| **3rd year undergraduate** | 1.12 | 0.70 | 1.53 | 0.817 |
| **4th year undergraduate** | 1.06 | 0.70 | 1.47 | 0.989 |
| **5th year or more** | 0.98 | 0.64 | 1.39 | 0.899 |
| **Enrollment Status (Ref = Full-time)** |  |  |  |  |
| **Other** | 0.83 | 0.51 | 1.66 | 0.755 |
| **Part-time** | 1.11 | 0.62 | 1.94 | 0.8 |

**Note.** Nagelkerke R² = .20; χ²(22, N = 798) = 78.56; *p < .05, **p < .001.

**Table S3**

Fully Adjusted Logistic Regression Predicting Low Perceived Restfulness (<3 Days per Week) Among U.S. College Students Aged 18–24 (N = 798)

| **Variable** | **Adjusted OR** | **95% CI Lower** | **95% CI Upper** | **p-value** |
| --- | --- | --- | --- | --- |
| **Psychological Distress** | 3.10 | 2.10 | 4.58 | < .001** |
| **Loneliness** | 1.85 | 1.25 | 2.74 | .002* |
| **Tobacco Use** | 1.40 | 0.95 | 2.05 | .080 |
| **Alcohol Use** | 1.70 | 1.15 | 2.50 | .007 |
| **Cannabis Use** | 1.55 | 1.05 | 2.28 | .025* |
| **Age (ref = 18)** |  |  |  |  |
| **19** | 0.91 | 0.65 | 1.78 | 0.834 |
| **20** | 0.84 | 0.66 | 1.48 | 0.956 |
| **21** | 0.99 | 0.55 | 1.61 | 0.837 |
| **22** | 0.98 | 0.54 | 1.42 | 0.717 |
| **23** | 0.76 | 0.54 | 1.30 | 0.585 |
| **24** | 0.80 | 0.72 | 1.50 | 0.73 |
| **Sex (ref = Female** |  |  |  |  |
| **Male** | 1.22 | 0.65 | 1.44 | 0.67 |
| **Non-binary** | 1.28 | 0.52 | 2.08 | 0.556 |
| **Race (Ref = White)** |  |  |  |  |
| **Asian or Asian American** | 0.91 | 0.58 | 1.56 | 0.698 |
| **Black or African American** | 0.98 | 0.70 | 1.70 | 0.812 |
| **Hispanic or Latino/a/x** | 1.18 | 0.61 | 1.61 | 0.659 |
| **Others** | 0.91 | 0.66 | 1.85 | 0.923 |
| **Year in School (Ref = 1 year undergraduate** |  |  |  |  |
| **2nd year undergraduate** | 0.95 | 0.68 | 1.28 | 0.729 |
| **3rd year undergraduate** | 1.02 | 0.59 | 1.66 | 0.802 |
| **4th year undergraduate** | 1.14 | 0.58 | 1.38 | 0.973 |
| **5th year or more** | 0.91 | 0.67 | 1.54 | 0.877 |
| **Enrollment Status (Ref = Full-time)** |  |  |  |  |
| **Other** | 0.96 | 0.47 | 1.85 | 0.785 |
| **Part-time** | 1.10 | 0.60 | 1.98 | 0.758 |

**Note.** Nagelkerke R² = .21; χ²(22, N = 798) = 83.44; *p < .05, **p < .001.
